# Anti SARS-CoV2 seroprevalence in Zanzibar in 2021 before the Omicron wave

**DOI:** 10.1101/2022.04.23.22274199

**Authors:** Salum Seif Salum, Mohammed Ali Sheikh, Antje Hebestreit, Sørge Kelm

## Abstract

**Objectives:** For Tanzania including Zanzibar, the development of the COVID-19 pandemic has remained unclear, since reporting cases was suspended during 2020/21. The present study provides first data on the COVID-19 seroprevalence among Zanzibari before the omicron variant wave starting in late 2021.

**Design:** During August through October 2021 representative cross-sectional data were collected from randomly selected households in 120 wards of the two main islands, Unguja and Pemba. Participants voluntarily provided blood samples to test their sera for antibodies against SARS-CoV2 in a semiquantitative enzyme-linked immunosorbent assay (ELISA).

**Results:** 58.9% of the 2051 sera analysed were positive without significant differences between Unguja and Pemba or between rural and urban areas, similar to observations from other sub-Saharan Africa countries.

**Conclusions:** The antibody levels observed are most likely to previous infections with SARS-CoV2, since vaccination was basically not available before the survey. Therefore, this study provides first insight, how many Zanzibari have had COVID-19 before the Omicron variant. Further, it provides the appropriate basis for a follow-up survey addressing how this seroprevalence influenced the susceptibility to the Omicron variants, given harmonised methodologies are used.

## Background

COVID-19 was declared as a global pandemic early 2020. Following first case reports in March (WHO, 2022), Tanzania implemented protective measures. However, May 2020 Tanzania stopped reporting COVID-19 cases to WHO and most restrictions were lifted 18^th^ May. In mid 2021 Tanzania resumed reporting, including previously detected COVID-19 cases, resulting in 33,836 confirmed cases and a death toll of 803 by 1 April 2022 (The United Republic of Tanzania, Ministry of Health, 2022). These data clearly showed COVID-19 waves for April-June 2020, January-March 2021, June-August 2021 coinciding with the original, alpha and delta variants waves, as well as for omicron variants, December 2021 through March 2022, (The United Republic of Tanzania, Ministry of Health, 2022).

Herd (or population) immunity is achieved, if a sufficient proportion of a population has acquired immunity, through natural infection or vaccination, to prevent further spread of the pathogen. To achieve this for COVID-19, vaccination is the most efficient and preferred way, according to WHO (WHO, 2020), although the proportion that must be vaccinated to effect herd immunity is still not known. While several efficient vaccines have been produced, in most sub-Saharan Africa countries these have only become available late and in low numbers. Zanzibar began vaccination July 2021 (Mikofu, 2021) and vaccination coverage in Tanzania including Zanzibar is still around 5% (The United Republic of Tanzania, Ministry of Health, 2022).

While confirmed COVID-19 cases correspond to only 0.5% of the population, an unknown number of SARS-CoV2 infections must be expected. Seroprevalence of anti-SARS-CoV2 antibodies can provide essential information (Bobrovitz *et al*., 2021). Therefore, this study was performed in 2021 providing first data of the COVID-19 seroprevalence among Zanzibari before the omicron variants wave started in November 2021.

## Methods

### Study area, study design and sampling

This cross-sectional study was designed to provide representative seroprevalence data from the main islands, Unguja and Pemba. 120 of the total 388 wards (Shehias) were randomly selected covering urban and rural areas. 354 randomly selected households were visited from 28^th^ July through 20^th^ October 2021. All household members were enrolled, irrespective of their age or sex, as in a previous survey (Nyangasa *et al*., 2016). All eligible participants gave written consent in an entirely voluntary manner after all relevant information had been provided in the local language.

Venous blood samples were collected into clotting activation tubes and transported to the laboratory for processing. Following centrifugation serum was stored at -80°C.

### Laboratory Analysis

The EUROIMMUN Anti-SARS-CoV-2 ELISA (IgG) kit provided a semi-quantitative analysis detecting human IgG antibodies against S1 domain of SARS-CoV-2 spike protein indicating prior infections and has been recommended for seroprevalence surveys like this (Gededzha *et al*., 2021). Using the calibrator of the kit, a ratio-based analysis (OD of sample/OD of calibrator) of the obtained data was performed to differentiate between negative (ratio below 0.8), positive (ratio above 1.1) and borderline cases. According to the manufacturer’s information, this kit displayed a positive agreement of 90% (95%CI=73.5%-97.9%), counting borderlines as negative, and a negative agreement of 100.0% (95%CI=95.5%-100.0%).

## Results and Discussion

Until now, the frequency of SARS-CoV-2 infections causing COVID-19 in Tanzania has been unknown. Seroprevalence provides important information how many individuals were exposed to SARS-CoV-2 before sampling. This study is the first report on anti-SARS-CoV-2 sero-prevalence in Zanzibar population. In total, 2,071 participants (66.2% of 3,143) from 349 (98.6%) households of this survey provided blood samples, from which 2,051 serum samples were available to be tested for anti-SARS-CoV-2 IgG; 56.9% of these samples were positive. Since vaccination against Covid-19 basically had not been available to participants, the antibodies detected are likely to be induced by preceding SARS-CoV-2 infection(s) during COVID-19 waves (The United Republic of Tanzania, Ministry of Health, 2022).

Seroprevalence was relatively evenly distributed and only minor differences were observed (Table 1) without significant differences (*P*>0.9999) between Unguja and Pemba. On Unguja, the seroprevalence observed in urban districts Magharibi and Mjini was lower, whereas on Pemba the lowest was in Micheweni, a rural district (Table 1). Prevalence was slightly higher in males compared to the female participants. The highest seroprevalence was observed in adolescents (*P*>0.9137) (Table 2).

**Table 1.** Regional seroprevalences; total number of tests conducted in Unguja and Pemba and their respective districts with seropositivity in percent.

| District | Seropositive<br>(N [%]) | Seronegative<br>(N [%]) | Borderline<br>(N [%]) | Total<br>(N) |
| --- | --- | --- | --- | --- |
| Total (Unguja) | 773 [57.1%] | 486 [35.9%] | 94 [6.9%] | 1353 |
| North A (Kaskazini A, Unguja) | 84 [56.8%] | 53 [35.8%] | 11 [7.4%] | 148 |
| North B (Kaskazini B, Unguja) | 93 [62.4%] | 49 [32.9%] | 7 [4.7%] | 149 |
| Central (Kati, Unguja) | 160 [58.4%] | 93 [33.9%] | 21 [7.7%] | 274 |
| South (Kusini, Unguja) | 81 [59.1%] | 50 [36.5%] | 6 [4.4%] | 137 |
| West (Magharibi, Unguja) | 159 [53.4%] | 122 [40.9%] | 17 [5.7%] | 298 |
| Urban (Mjini, Unguja) | 196 [56.5%] | 119 [34.3%] | 32 [9.2%] | 347 |
| Total (Pemba) | 394 [56.4%] | 267 [38.3%] | 37 [5.3%] | 698 |
| Chake (Pemba) | 112 [60.5%] | 59 [31.9%] | 14 [7.6%] | 185 |
| Micheweni (Pemba) | 85 [50.6%] | 74 [44.0%] | 9 [5.4%] | 168 |
| Mkoani (Pemba) | 127 [56.2%] | 89 [39.4%] | 10 [4.4%] | 226 |
| Wete (Pemba) | 70 [58.8%] | 45 [37.8%] | 4 [3.4%] | 119 |
| Total (Zanzibar) | 1167 [56.9%] | 753 [36.7%] | 131 [6.4%] | 2051 |

**Table 2.** Sex and age distribution of seroprevalences; total number and percentage of tests conducted.

| Population tested | Seropositive<br>(N [%]) | Seronegative<br>(N [%]) | Borderline<br>(N [%]) | Total<br>(N) |
| --- | --- | --- | --- | --- |
| Female | 626 [56.1%] | 416 [37.3%] | 74 [6.6%] | 1116 |
| Male | 541 [57.9%] | 337 [36.0%] | 57 [6.1%] | 935 |
| age group <10 years | 152 [51.9%] | 125 [42.7%] | 16 [5.5%] | 293 |
| age group 10 - <14 years | 173 [61.6%] | 97 [34.8%] | 10 [3.6%] | 280 |
| age group 14 - <17 years | 121 [66.5%] | 49 [26.9%] | 12 [6.6%] | 182 |
| age group 17 - <60 years | 601 [54.9%] | 412 [37.7%] | 81 [7.4%] | 1094 |
| age group >60 years | 120 [59.4%] | 70 [34.7%] | 12 [5.9%] | 202 |
| Total participants | 1167 [56.9%] | 753 [36.7%] | 131 [6.4%] | 2051 |

In summary, this first population-based survey uncovered widespread SARS-CoV-2 seropositivity across all districts of Zanzibar prior to the onset of the current Omicron-dominated wave. Overall, seroprevalences observed are similar to those observed in other sub-Sahara Africa countries (Lewis *et al*., 2022).

At present, this level of seroprevalence cannot be considered as herd immunity, since it is uncertain, how antibody levels correlate with virus neutralisation, protection against re-infection, symptomatic and asymptomatic cases (Centers for Disease Control and Prevention, 2020, Hamady *et al*., 2022). In this respect, comparing the seroprevalences reported here with the current situation after/during the Omicron variants wave could provide important information, if using identical tools complementing the analysis with neutralising activities against Omicron and previous variants in these sera.

## Data Availability

All data produced in the present work are contained in the manuscript

## Ethical approval statement

The study has been carried out in accordance with The Code of Ethics of the World Medical Association (Declaration of Helsinki) for experiments involving humans and was approved by The Second Vice President Office and The Ministry of Health Zanzibar through the Zanzibar Medical Research and Ethics Committee (ZAMREC/0001/AUGUST/013).

## Acknowledgement

Authors are grateful to all participants for their invaluable contribution as well as to all Shehas of the participating wards for their the support. We highly appreciate the restless technical support by Petra Berger and all members of the Unguja and Pemba survey teams. The authors acknowledge very much the efforts of the Public Health Laboratory, Pemba, the Zanzibar Health Research Institute, and the Ministry of Health for their essential support, which they devoted for successful completion of this study.

---

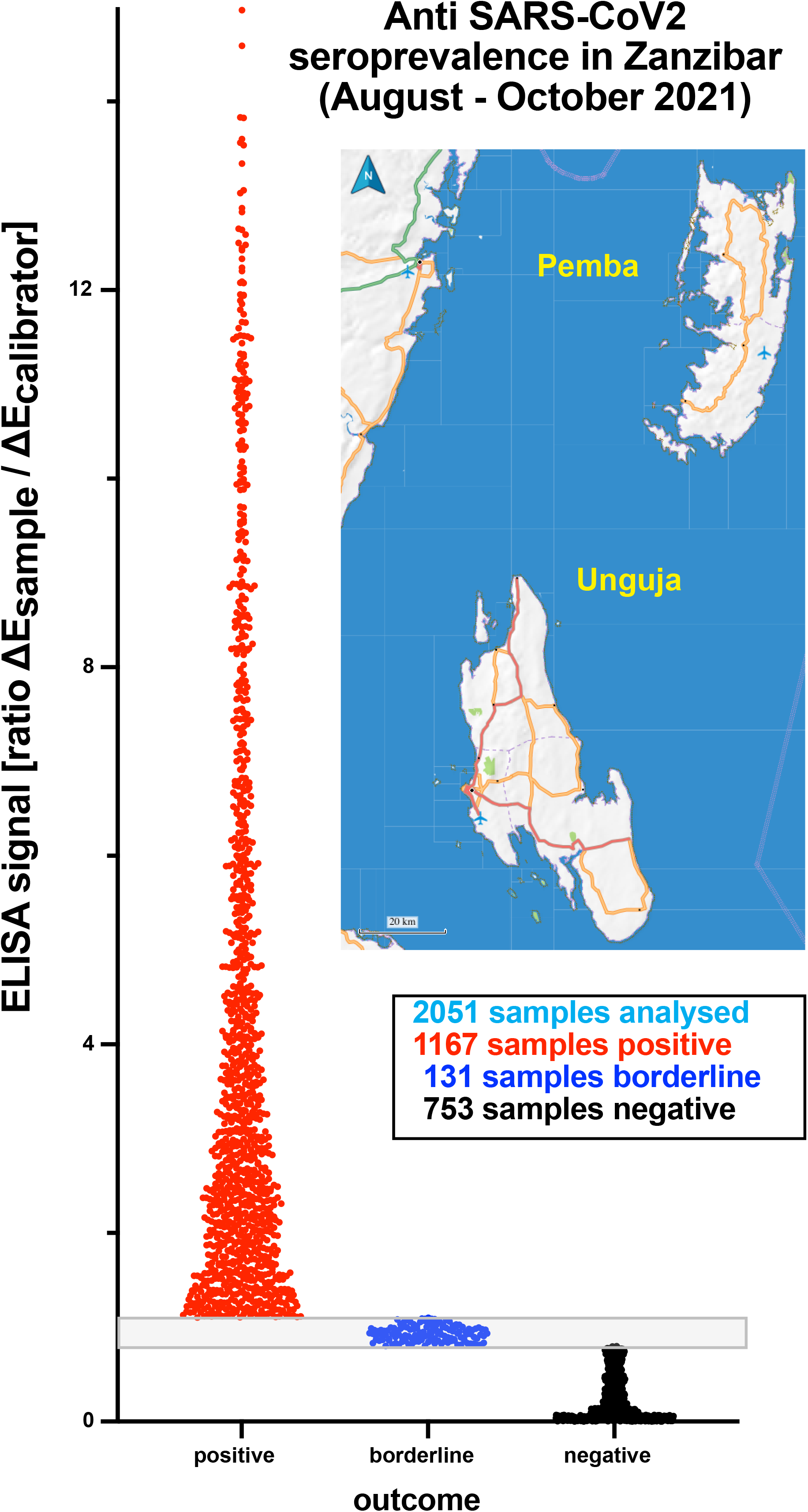

## References

Bobrovitz N, Arora RK, Cao C, Boucher E, Liu M, Donnici C, et al. Global seroprevalence of SARS-CoV-2 antibodies: A systematic review and meta-analysis. PLoS ONE 2021; 16(6): e0252617. https://doi.org/10.1371/journal.pone.0252617

Centers for Disease Control and Prevention. What COVID-19 Seroprevalence Surveys Can Tell Us. https://www.cdc.gov/coronavirus/2019-ncov/covid-data/seropreval-ance-surveys-tell-us.html, 2020 (visited 18.04.2022)

Gededzha MP, Mampeule N, Jugwanth S, Zwane N, David A, Burgers WA, et al. Performance of the EUROIMMUN Anti-SARS-CoV-2 ELISA Assay for detection of IgA and IgG antibodies in South Africa. PLoS ONE 2021; 16(6): e0252317. https://doi.org/10.1371/journal.pone.0252317

Hamady, A., Lee, J., Loboda, Z.A. Waning antibody responses in COVID-19: what can we learn from the analysis of other coronaviruses? Infection 2022; 50, 11–25. https://doi.org/10.1007/s15010-021-01664-z

Lewis HC, Ware H, Whelan M, Subissi L, Li Z, Ma X, et al. SARS-CoV-2 infection in Africa: A systematic review and meta-analysis of standardised seroprevalence studies, from January 2020 to December 2021; medRχiv preprint 2022; https://doi.org/10.1101/2022.02.14.22270934 (visited 18.04.2022)

Mikofu J. Zanzibar kicks of Covid-19 vaccination, orders another consignment. The Citizen, 16 July 2021, https://allafrica.com/stories/202107160932.html, 2021 (accessed 18 April 2022)

Nyangasa MA, Kelm S, Sheikh MA. Hebestreit A. Design, response rates, and population: characteristics of a cross-sectional study in Zanzibar, Tanzania. JMIR Res Protoc. 2016; 5(4):e235. https://doi.org/10.2196/resprot.6621

The United Republic of Tanzania, Ministry of Health, Covid-19 Situation Report No 29; https://www.moh.go.tz/storage/app/uploads/public/625/527/243/625527243c3bd391718416.pdf, 2022 (visited 18.04.2022)

WHO, Coronavirus disease (COVID-19): Herd immunity, lockdowns and COVID-19, WHO,31 Dec 2020; https://www.who.int/news-room/questions-and-answers/item/herd-immunity-lockdowns-and-covid-19, 2020 (accessed 18 April 2022)

WHO, Coronavirus (COVID-19) Dashboard. https://covid19.who.int/region/afro/country/tz, 2022 (accessed 18 April 2022)

